# An organising framework for healthcare decarbonisation research: an exploratory classification study

**DOI:** 10.1101/2025.11.21.25340730

**Authors:** Mila Petrova, Fanny Burrows, Jan W. van der Scheer, Timoleon Kipouros, James Smith

## Abstract

**Objectives:** To develop an organising framework for healthcare decarbonisation research which goes beyond classification schemes based on Scope 1, 2 and 3 emissions or lists of loosely connected themes, and which is intended to support the coordination, funding and application of research into policy and practice. The organising framework should be focused on the NHS in England but enable application to healthcare systems more broadly.

**Design:** An exploratory manual (non-machine-led) classification study of over 160 research questions derived from a scoping review of 10 systematic reviews (118 screened), 13 stakeholder documents (35 key stakeholder websites searched), two research priority exercises, and four research funder sources (over 430 funding areas screened). The above and a further 21 sources were also used to identify areas without explicit research questions but of clear thematic relevance.

**Setting:** Primarily high-income healthcare systems, with a focus on the NHS in England.

**Participants:** Not applicable.

**Primary outcome:** A multi-level thematic framework representing current and missing areas of research in healthcare decarbonisation.

**Results:** The framework comprises six top-level themes, grouping 39 sub-themes at level two, and 86 sub-themes at level three. The top-level themes are: **N**atural resource use and sources of carbon; **H**ealthcare contexts; **S**olutions; **S**takeholders; **O**rganisational levers for change; and **S**cientific measurement and theory (the “NHS-SOS framework”).

**Conclusions:** This framework offers a structured, empirically derived representation of the emerging field of healthcare decarbonisation research. It is intended as a living tool to support shared understanding, prioritisation and action, and to foster coherence in a currently fragmented research landscape.

**Article summary:** *Strengths and limitations of this study:*

- The study used a transparent and structured process to derive themes from over 160 research questions, sourced from a diverse set of systematic reviews, stakeholder documents, research priority exercises, and funding calls.
- The inductive approach respected the complexity, breadth and multiple perspectives inherent to healthcare decarbonisation research.
- The study drew on a wide range of sources selected for conceptual and perspectival breadth but was nonetheless small relative to the volume of publications in the field.
- Many of the research questions were not explicitly stated in the source documents and had to be derived through interpretive analysis. This introduced a potential for bias, which was mitigated through a clearly documented and transparent process outlining how interpretations were made.

## INTRODUCTION

Healthcare systems are major contributors to global greenhouse gas (GHG) emissions, accounting on average for an estimated 4.9% of national emissions, with a reported range of 1.5% to 9.8%.[1] If healthcare were a country, it would rank as the fifth largest emitter on the planet.[2] Recognising the urgency of addressing this impact, the World Health Organization (WHO) and its member states have called for action “at pace and scale”, leading to the formation of the Alliance for Transformative Action on Climate and Health (ATACH).[3] The National Health Service (NHS) in England has taken a leading role in healthcare decarbonisation by setting ambitious targets to reach net zero by 2040 for its direct footprint and 2045 for its extended footprint, including that of its supply chain.[4] A 2024 systematic review found that among six countries with available time series data, only NHS England and NHS Scotland had demonstrably reduced their healthcare-related emissions.[1] While peer-reviewed literature may not fully reflect the progress of many countries, the progress made by the NHS stands out as sector-leading.

Decarbonisation of healthcare is a complex, system-wide challenge involving a wide array of stakeholders, including government bodies, healthcare commissioners and providers, non-profit organisations (NGOs), businesses, professional communities, and patients. The research community has a critical role in supporting healthcare decarbonisation efforts by generating the evidence to inform action through policy and practice. Both decarbonisation activities, and the research which supports them, have typically been organised either by emission sources or broad thematic areas without a unifying structure. Examples of organisation by emission sources include Greenhouse Gas Protocol’s three emission scopes [5,6] and the NHS Carbon Footprint and Carbon Footprint Plus used in England.[7] Examples of organisation by broad themes include the nine inductively generated themes in Braithwaite and al.’s comprehensive systematic review of 205 studies on strategies to decarbonise healthcare [8] and the 19 themes of the Resource Library of the Centre for Sustainable Healthcare.[9] Controlled vocabularies and formal ontologies for databases have also made little headway. For instance, the Medical Subject Headings (MeSH) thesaurus of the National Library of Medicine, which represents the fields of biomedicine and health for the purposes of cataloguing information, does not include the term “decarbonization” and only includes three concepts in its tree for Climate Change.[10]

Without a broadly shared representation of the field, the research community risks fragmented efforts, missed synergies, and overlooked gaps. A common frame of reference – here termed an ‘organising framework’ – could enable more systematic identification of research needs, support system-level, coordinated research and action across programmes of work, and facilitate knowledge exchange. This could be of particular importance as new evidence is generated and knowledge gaps are filled. While a fully standardised, finalised consensual map of the field may be neither feasible nor desirable, a shared, evolving framework can offer structure to key efforts such as capturing and synchronising practical evidence-based action, guiding impactful research, informing debate, and forging new links between currently disconnected or weakly connected programmes and stakeholders. It may also support greater consensus on how research in this area is conceptualised, organised and reported, helping to build a more coherent and cumulative evidence base.

The aim of this work was to develop an organising framework for healthcare decarbonisation research, focused on the NHS in England but applicable to healthcare systems more broadly, and intended to support the coordination, funding and application of research into policy and practice.

## METHODS

We developed an organising framework for healthcare decarbonisation research by abstracting shared features of and classifying research questions identified through a scoping review of relevant systematic reviews, stakeholder documents, research priority exercises and funding calls. A distinctive feature of our approach was the highly transparent process of identification and formulation of research questions, including many that could not be directly extracted from the source documents but were derived through an interpretive process necessitating subjective judgements, which we aimed to render as auditable as possible. “Transformations” of the contents of source documents to derive research questions from them were often necessary where documents addressed broader, narrower or different contexts than our focus, namely the UK healthcare system (NHS). Further detail on our methodological approach is provided below.

### Source documents

The pool of source documents was selected to maximise conceptual breadth across both research and policy domains (see **Supplement 1** for details on the selection process and a list of sources). 10 systematic reviews (out of 118 screened), 13 stakeholder documents (35 key stakeholder websites searched), two research priority exercises, and four research funder sources (out of over 430 funding themes, calls and associated projects screened) were used to derive research questions.

### Research questions

The process by which research questions were derived from the source documents is described in detail in **Supplement 2** and is summarised below. Seven types of source contents were used to extract (word-by-word) or formulate (through an interpretative process) research questions:

(1) research gaps or recommendations,
(2) research limitations,
(3) research availability,
(4) research-policy-practice collaborations,
(5) intentions to fund or commission research,
(6) recommendations for guideline development, and
(7) commitments or requirements for practical action (e.g. policy commitments or directives) that implied a need for new, specialised knowledge.

Where possible, contents of these seven types were used to directly extract research questions. In many cases, however, research questions had to be formulated from the source contents through an interpretative process that applied one or more of the following eight transformations:

(1) attributing a focus on decarbonisation,
(2) (re-)specifying boundaries (the context to which the research question pertains),
(3) splitting or combining in new ways elements from complex statements,
(4) raising the level of abstraction,
(5) articulating further concepts or contents,
(6) adding a research perspective,
(7) adding a meta-science perspective, and
(8) editing for standalone clarity.

### Approach to developing the organising framework

Research questions were grouped together and/or a category (class, theme) was formed and labelled by identifying and abstracting the core characteristic(s) of one or more questions.

Often, the process of comparing questions to determine lines of similarity and difference in order to group them together or keep them apart coincided with the process of forming and labelling a category. Categories could, however, be also formulated from single questions. In some cases, we also added ‘placeholder themes’ to the framework which did not have a corresponding research question. Placeholder themes were generated in two ways. Some reflected areas of limited knowledge highlighted in source documents whose content was not specific enough to support a well-formulated research question. Others corresponded to a research gap identified in dated sources which we read as background information due to their high relevance to the topic. In both cases, the placeholder theme captured a conceptual gap in the evolving organising framework. Placeholder themes are clearly labelled as such to indicate that a theme may not currently reflect a robust research need, due to the limited or outdated nature of the underlying data.

The above process was anchored in ideas from ontology development in information science and philosophical frameworks such as promiscuous realism and pluralistic realism.[11–14] In information science, ontologies provide a shared vocabulary and structure for organising domain knowledge, making assumptions explicit, and enabling sharing, reuse, integration and comparison of domain knowledge. Philosophically, promiscuous and pluralistic realism acknowledge that while an objective reality exists, there are multiple valid ways to interpret and classify entities and phenomena within it. These philosophical concepts informed much of our approach to labelling and grouping themes, recognising the value of representing multiple perspectives and avoiding overly rigid or reductionist structures.

A set of further principles guided our approach to labelling and grouping themes based on research questions and developing the overall structure of the framework (in discussing the framework below, we will prioritise the term “themes” as opposed to “categories” or “classes”). Altogether, these principles were:

(1) taking multiple perspectives,
(2) giving weight to the mental models of policy makers,
(3) revising dynamically while allowing for periods of stabilisation,
(4) accommodating variable levels of commitment to decarbonisation, including resistance to it,
(5) avoiding technical language and rigid hierarchies when they conflict with intuitive use,
(6) incorporating opportunities to align with external conceptual models,
(7) attending to conflicts and discrepancies,
(8) balancing transferability with domain specificity,
(9) co-locating topics to enable syntheses rather than reinforcing silos,
(10) accommodating blurred boundaries between decarbonisation, climate change and sustainability.

Further detail on each principle is provided in **Supplement 2**. Derivation of research questions, abstraction of themes from them, and restructuring of the framework continued until the latter reached a point of stability, at which no major new themes or structural revisions were introduced.

### Patient and public involvement

There was no direct patient and public involvement in this study. It is recognised that patients are key stakeholders in the healthcare system and as such their input will be sought in future developments of the framework.

## RESULTS

We derived over 160 research questions from across 10 systematic reviews, 13 stakeholder documents, two research priority exercises, and four research funder sources. The process of classifying these research questions, using 10 key principles, resulted in the proposed organising framework for healthcare decarbonisation research: the NHS-SOS framework, as per the initials of the six top-level themes:

- **N**atural resource use and sources of carbon
- **H**ealthcare contexts
- **S**olutions
- **S**takeholders
- **O**rganisational levers for change
- **S**cientific measurement and theory

These six ‘level 1’ themes (represented in **Figure 1**) include 39 more specific ‘level two’ sub-themes (**Table 1**), with the latter broken down further into 86, even more granular, ‘level three’ sub-themes. A small number of ‘level four’ sub-themes were also generated. The full framework, including level three and four sub-themes, is presented in **Supplement 3**.

**Figure 1:**
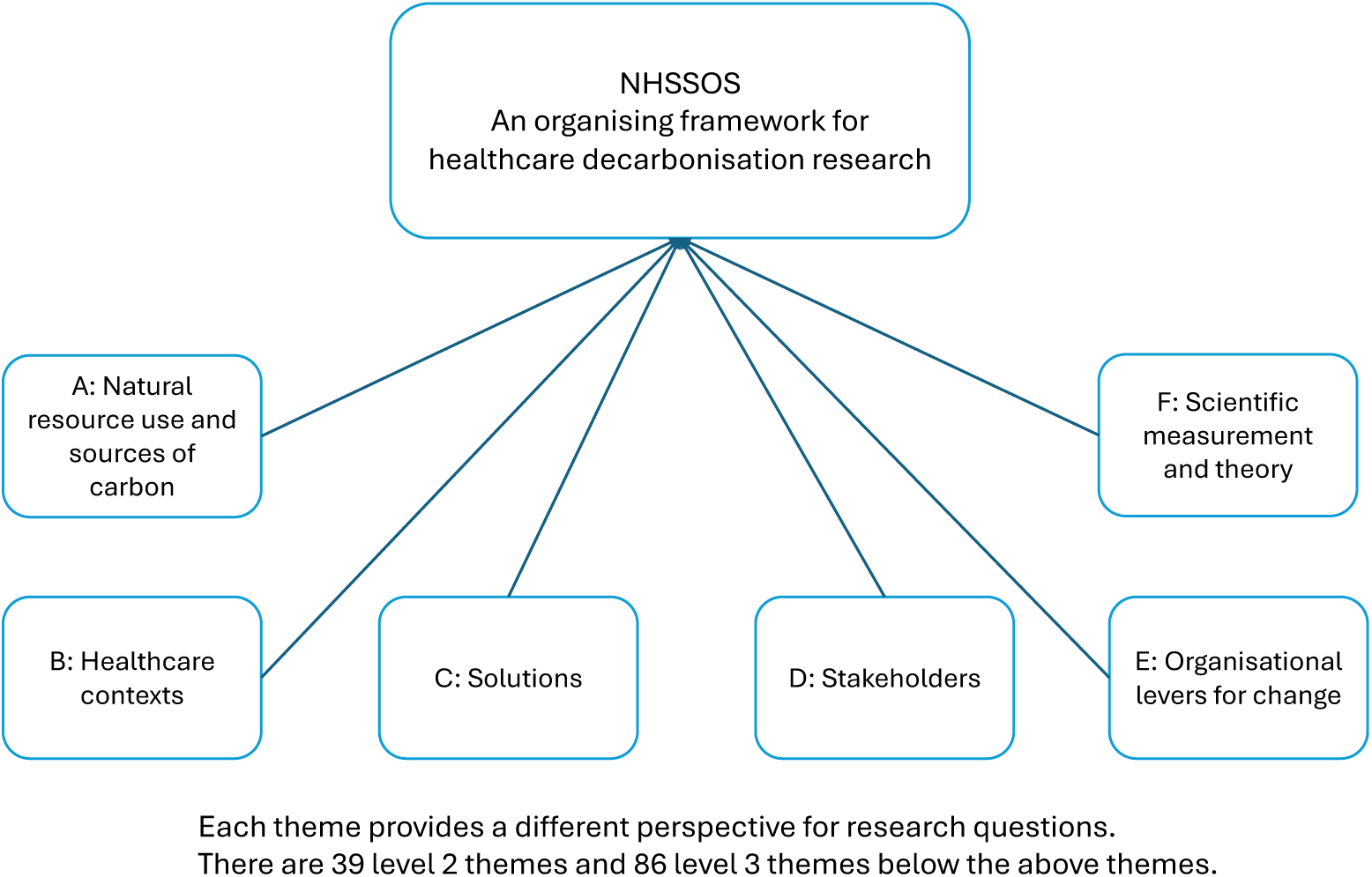
Organising framework for healthcare decarbonisation research (NHS-SOS) with top-level (‘level 1’) themes

**Table 1:** Organising framework for healthcare decarbonisation research (NHS-SOS) with level 1 and 2 themes.

|  |
| --- |
| **A. NATURAL RESOURCE USE AND SOURCES OF CARBON** |
| A1. Energy sources and water use in healthcare |
| A2. Medicines, equipment and consumables as decarbonisation targets; associated production, procurement and supply chains |
| A3. Waste, reuse, recycling and circularity in healthcare |
| A4. Food and nutrition in healthcare contexts and for health-related outcomes |
| A5. Transport, travel and mobility for healthcare purposes and in healthcare contexts |
| **B. HEALTHCARE CONTEXTS** |
| B1. Decarbonisation of healthcare spaces (buildings, facilities, estates, sites, etc.) |
| B2. Decarbonisation of healthcare settings (primary care, secondary care, community care, etc.) [PLACEHOLDER] |
| B3. Decarbonisation of clinical specialities and/or health conditions |
| B4. Decarbonisation of healthcare supply chains |
| B5. Sustainable reorganisation of healthcare systems in contexts of emergencies, disasters and other stresses and shocks |
| **C. SOLUTIONS** |
| C1. Solutions for decarbonising healthcare (big picture, cross-cutting issues) |
| C2. Digital solutions |
| C2. Innovative materials |
| C3. Innovative designs |
| C4. Capture and removal of greenhouse gases [PLACEHOLDER] |
| C5. New models of care and decarbonisation |
| C6. Combining innovation and 'back to basics' approaches |
| C7. Improved healthcare efficiency as a route to decarbonisation |
| C8. Education, training and information provision |
| C9. Behaviour change-focused solutions for decarbonising healthcare |
| C10. Complex interventions for decarbonising healthcare |
| C11. The carbon footprint of adaptations to climate change and decarbonisation solutions themselves |
| **D. STAKEHOLDERS** |
| D1. Stakeholders in healthcare and/or decarbonisation (big picture, cross-cutting issues) |
| D2. The healthcare workforce and the decarbonisation of healthcare |
| D3. Types of healthcare workers and the decarbonisation of healthcare |
| D4. Patient populations and groups and the decarbonisation of healthcare [PLACEHOLDER] |
| **E. ORGANISATIONAL LEVERS FOR CHANGE** |
| E1. Policy and governance for the decarbonisation of healthcare |
| E2. Funding and financial mechanisms for the decarbonisation of healthcare |
| E3. Practical guidance for the decarbonisation of healthcare |
| E4. Certification and accreditation schemes for healthcare decarbonisation [PLACEHOLDER] |
| E5. Visions, models and frameworks of healthcare provision which incorporate decarbonisation principles |
| E6. Networks, communities of practice and platforms for collaborative work on healthcare decarbonisation |
| E7. Managing (perceived) trade-offs and conflicts of values in decarbonising healthcare |
| ***F. SCIENTIFIC MEASUREMENT AND THEORY*** |
| F1. Measuring the carbon footprint and the decarbonisation of healthcare |
| F2. Concepts, frameworks and theories concerning the decarbonisation of healthcare |
| F3. Healthcare decarbonisation in the social conversation |
| F4. Needs for quality improvement in research on healthcare decarbonisation |
| F5. Systematic approaches to identifying research and gaps in research on healthcare decarbonisation |
| F6. Turning knowledge into action on healthcare decarbonisation |

## DISCUSSION

### Main findings

We developed an organising framework for healthcare decarbonisation research (NHS-SOS) based on research questions derived from systematic reviews, stakeholder documents, research priority exercises and research funder sources. The framework comprises six top-level themes, each representing a distinct perspective for examining research questions about healthcare decarbonisation, supported by three further levels of sub-themes. These six themes function as both lenses for inquiry and domains of action. The framework provides a starting point for constructing a shared representation of the field of healthcare decarbonisation to enable systems thinking and systemic action.

### Study strengths and limitations

To our knowledge, this is the first detailed, multi-level mapping of the field of healthcare decarbonisation research. The inductive approach underpinning the generation of themes captured how researchers, policy makers, and practitioners are engaging with the challenge of decarbonisation. It respected the complexity, breadth and multi-perspectival nature of the landscape of healthcare decarbonisation research and avoided imposing narrow conceptualisations, e.g. solely carbon-footprint based, or unrealistically rational pre-existing ones. The logically disciplined, even if flexible and user-friendly structure, in turn, avoided the downsides of lists of loosely connected themes, often combining themes of different levels of generality and using different lenses.

The principles used in individuating, labelling and classifying themes were highly unusual as a combination but, we argue, have significant heuristic potential when jointly applied. They bring together ideas coming from the philosophy of science, particularly theories of classification; basic rules for designing domain ontologies; attention to the needs of policy makers; and a commitment to user friendliness, amongst other conceptual influences. These principles and their combination do, however, entail value judgements which merit further debate and articulation in the process of future co-production work on the organising framework.

While the framework was developed with a focus on the NHS in England, its research questions – and the themes and sub-themes they informed – are likely broadly applicable across healthcare system levels and transferable to healthcare systems in other countries, particularly higher-income ones. In addition, the method developed in this work for building an organising framework may hold relevance for other complex systemic challenges in healthcare research and beyond.

The data source of research questions on which the framework was based was obtained through a highly transparent and structured process from a diverse set of strategically selected sources. Nevertheless, it may also have biases arising from the limited number of included sources and the fact that many research questions could not be directly extracted but were formulated through interpretive processes. It is also important to acknowledge that there is a level of tension between the nature of the data source we used – research questions indicating research gaps – and the goal of representing a field of research. While research gaps signal strongly what has been researched, a process of mapping and classifying existing research may have generated a different outcome.

### Implications for research and practice

We envisage this framework being used in organising co-production processes around the prioritisation, design, conduct, dissemination and uptake of research to advance the decarbonisation of the NHS in England. With appropriate adaptation, the framework may also meet research-into-action needs of other healthcare systems, particularly in high-income countries.

A fully comprehensive framework – based on an exhaustive review of documents available in electronic databases and grey literature – was beyond the scope of this study but may also not be the most effective way forward. The field of healthcare decarbonisation is rapidly evolving while there is no shared representation of it. Sharing an initial framework now, intended to serve as a living tool for continued co-creation processes, is likely of more value than a fully polished framework later – not least given the urgency of the climate crisis.

Further development of the framework will require new empirical inputs and engagement and involvement processes. New empirical inputs may include not only additional systematic reviews and stakeholder documents, but also content which is unconventional in academic research. For instance, submissions to start-up business competitions and award schemes in the climate innovation space may highlight emerging, cutting-edge research and implementation areas of work. The analysis of ‘green plans’ of healthcare organisations and of project evaluations conducted in healthcare settings may offer themes grounded in operational realities.

Growing and refining the organising framework will also benefit from engaging with research and expertise concerning its formal structure – such as developments in classification theory and domain ontologies, and user experience research on how people interact with classification or organising systems.

Importantly, we see the ongoing development of the NHS-SOS framework as inseparable from the practical work of enabling system-level action on the decarbonisation of healthcare. The framework is not intended primarily as a theoretical model or cognitive heuristic, but as a practical tool to support a growing collective understanding of the field, foster systems thinking, and enable coordinated and contextualised action. It is envisaged as one tool, out of many, needed to improve the flow of research evidence into practice and policy in the field of decarbonisation and to ensure that practice and policy needs shape new research. We invite stakeholders to contribute to an open discussion about what a new generation of research-into-action infrastructure for healthcare decarbonisation should look like.

## CONCLUSION

This work presents an organising framework for healthcare decarbonisation research, developed through a rigorous inductive process and designed to accommodate multiple perspectives and points of entry into the field. The framework offers a foundation for structuring this emerging field and supporting the co-production of research infrastructure needed to accelerate research in response to the climate emergency.

## Supporting information

see Supplement 1 for details

## Data availability statement

The data that underlies the framework development are available on reasonable request from the corresponding author. Access to data may be granted to bona fide researchers under a data sharing agreement.

## Author Contributions

All authors contributed to the conceptualisation. MP undertook the data collection, method development, and analysis. MP and JS drafted the manuscript with review and editing from FB, JWvdS and TK. JS administered the project. JS is the guarantor for this article. All authors read and approved the final manuscript.

## Legends of figures, tables and supplements

**Supplement 1**: Identification and selection of sources; list of sources used

**Supplement 2**: Analysis approach – content types, transformations, principles of development

**Supplement 3:** Full NHS-SOS framework

