## Supplementary material for "An organising framework for healthcare decarbonisation research: an exploratory classification study": see Supplement 1 for details

### Supplement 1: Identification and selection of sources; list of sources used

UNDER REVIEW

This supplement first describes the process of identifying and selecting sources (Section A) and then lists the sources used (Section B).

#### *A. IDENTIFICATION AND SELECTION OF SOURCES*

***Process of narrowing down of preferred sources and forms of research needs***

Early discussions within the team revealed a variety of conceptualisations of the guiding concept of 'research needs'. Drawing on them and further analytical work by the first author (MP), we generated a working framework of 'forms and sources' of research needs, articulating 12 such forms and sources and over 20 approaches to operationalising the work. We then prioritised six of the forms and sources of research needs and associated approaches to identifying them. After scoping the work, we settled on a process which combined extraction/ derivation of research needs from:

1) **Systematic reviews** concerning health systems decarbonisation retrieved through a pragmatic, highly focused search strategy (see below). These were expected to have identified research gaps through a robust process of locating, mapping, critically evaluating and synthesising already available research.

2) **Key stakeholder sources** – flagship reports or other sources produced by key stakeholders working at the intersection between health and climate change. Such sources were expected to capture 1) knowledge gaps as experienced by a variety of non-academic stakeholders (which may be actual gaps in academic research or indicators of inefficiencies in knowledge dissemination and translation) and 2) knowledge gaps as revealed by the challenges of implementing decarbonisation practices, with the complexity of 'the frontline' and 'the whole' of the health system less likely to be present in academic sources.

**2a) Funding calls and recently funded projects** were particular subtypes of key stakeholder sources of interest. They were expected to capture a perspective on knowledge gaps that incorporates a cost-benefit analysis. Information on funded projects was also expected to signpost to both research "in the pipeline", which may soon fill in knowledge gaps identified through the other approaches, and highly innovative research which may be making inroads into the "unknown unknowns" of health system decarbonisations.

3) **Pre-existing lists of research needs**, as generated by other initiatives on identifying research/ knowledge gaps and developing research agendas.

***Process of identifying sources***

*Systematic reviews*

Systematic reviews concerning health systems decarbonisation were retrieved through a pragmatic, highly focused search strategy in MEDLINE, PubMed interface:

("net zero" OR "net 0" OR decarboni* OR sustainab* [ti] OR "climate change" [ti]) AND ("Health Policy"[MAJR] OR "health system" OR healthcare OR "health care")

The goal was to remain at the 'big picture' level (hence the keywords are very broad) and to reduce false positives (hence concepts broader than decarbonisation were only searched for in title and "health policy", aiming to capture research-policy interactions, was only included as a "major heading").

Searches were run in Jun 2024.

At the level of searches, no year, language or country limits were set. To limit to systematic reviews, we applied the embedded PubMed systematic reviews filter. Articles were considered eligible for inclusion if they represented systematic reviews on decarbonisation/ sustainability/ climate change issues at the level of health systems as a whole or at the level of individual components (namely settings, such as hospitals or primary care, or the WHO health system "building blocks", such as the healthcare workforce or governance). Articles from healthcare systems of high-income countries were prioritised due to the greater similarity between those

UNDER REVIEW

systems and the NHS. Screening of articles at title and abstract level was performed by one analyst (MP). Systematic reviews in reference lists or recommended by colleagues were also considered for inclusion.

5,290 publications were retrieved through the search strategy above, of which 118 were systematic reviews. Of these, 24 reviews were selected for further consideration following title and abstract screening. Three further reviews were identified in alternative ways (colleagues' recommendations or reference lists), for an initial shortlist of 27 reviews.

Ultimately, 10 reviews were selected for detailed data extraction (see list of sources below). Eight were used as background sources (2 of which were dated, respectively from 2009 and 2014, and 6 expanded the organising framework in ways that were fully expected conceptually, in 5 cases towards specific medical specialties and in 1 case towards a specific type of medicinal product). As with other background sources, these were used to test the emerging organising framework and to expand it through placeholder themes. ('Placeholder themes' were themes which were not illustrated by a specific research question, either because there was not enough information to generate a research question from the respective source or because the source was dated; further detail in the manuscript.). 9 reviews were excluded because their topic was already covered by an alternative source. At this exploratory stage, comparing the research recommendations of sources with a significant overlap of coverage was considered low priority. In 7 cases (systematic reviews on surgery and operating theatres), the alternative source was the *James Lind Alliance* list [11]. The other two topics for which we had chosen an alternative source were nutrition and digital health.

*Key stakeholder sources*

A list of 30+ key stakeholders was generated and iteratively enriched through suggestions of team members. The key stakeholders were organisations and initiatives which are either:

- firmly positioned at the intersection between healthcare and decarbonisation/ climate change/ environmental sustainability (Type 1 stakeholders), or
- have an influential role in healthcare improvement and can thus be expected to lead and/or enable healthcare decarbonisation (Type 2 stakeholders).

UNDER REVIEW

Type 1 stakeholders were primarily UK-based organisations placed in the context of major global actors. Being based in the UK did not, however, seem to be associated with a primary focus on the UK for many of the selected stakeholders, by virtue of climate change being a planetary issue. Type 2 stakeholders were only from the UK.

**Box S1a: Key actors, type 1: Organisations with a primary remit at the intersection between healthcare & decarbonisation/ sustainability/ climate change (UK in global context)**

**UK-centred**

**Health system structures** (for health system structures for Wales and Northern Ireland, see 16 and 17, Box S1b)
1. Greener NHS (NHS England)
2. NHS Sustainability Action (NHS Scotland)

**Other country-level structures** (not part of the health system governance structure)
3. Green Health Wales
4. Climate Northern Ireland, Health & Wellbeing sector

**Non-profit organisations**
5. Centre for Sustainable Healthcare
6. UK Health Alliance on Climate Change
7. Sustainable Healthcare Coalition
8. Greener Practice: UK's Primary Care Sustainability Network

**Business-oriented networks**
9. Climate and Health Coalition (Forum for the Future)

**Advocacy and pressure groups**
10. Health for Extinction Rebellion (UK)

**Global**
11. Alliance for Transformative Action on Climate and Health (ATACH)
12. Health Care Without Harm
13. Health and Environment Alliance (HEAL)
14. The Global Climate & Health Alliance

**Flagship commissions (UK or global) at the intersection between healthcare & decarbonisation/ sustainability/ climate change**

15. Lancet Commission on Sustainable Healthcare

UNDER REVIEW

#### Box S1b: Key actors, type 2: UK organisations driving healthcare quality and improvement (UK only)

**Health system structures** (for England and Scotland, see 2a)

16. NHS Wales
17. Health and Social Care Northern Ireland

**Organisations responsible for setting and/or monitoring compliance with healthcare standards**

18. National Institute for Health and Care Excellence (NICE)
19. Care Quality Commission

**Networks creating links between the NHS and health research and innovation**

20. Health Innovation Network (HIN)
21. National Institute for Health and Care Research Applied Research Collaborations (NIHR ARCs)
22. National Institute for Health and Care Research HealthTech Research Centres (NIHR HRCs)

**Think-tanks with a health sector focus**

23. The Health Foundation – done, but perhaps leave out for consistency
24. The King's Fund
25. The Nuffield Trust

**Communities of practice for healthcare quality and improvement**

26. The Academy of Fabulous NHS Stuff

**Patient and public involvement organisations**

27. Healthwatch
28. The Patients Association
29. NHS PPI groups

**Major UK funders with a remit in health**

30. National Institute for Health and Care Research (NIHR)
31. The Wellcome Trust
32. UK Research and Innovation (UKRI)
33. Advanced Research + Invention Agency (ARIA)
34. Association of Medical Research Charities (AMRC)
35. NHS Charities Together

**Other organisations that have produced research-intense documents at the intersection between healthcare & decarbonisation/ sustainability/ climate change were opportunistically identified.**

UNDER REVIEW

The preferred type of key stakeholder source was a flagship document (report, paper, strategy, roadmap, etc.) produced or co-produced by a particular organisation, addressing the decarbonisation of healthcare overall or of the part of the healthcare system that falls within the remit of the organisation (e.g. primary care).

If such a document was not available, any flagship document at the intersection between health/ healthcare & climate change/ the environment was considered. Ideally, the document also dedicated focused attention to the research needed in the domain or had a substantial research underpinning (e.g. was based on an extensive literature review). An exemplar document in terms of attention to research needs (though not a thematic exemplar) was the "Heat Resilience Strategy" of the Physiological Society and Faculty of Public Health [25].

Documents were primarily sought in "Publications" or similar sections of organisational websites. If there was no such section, website pages were reviewed until a document which met the above criteria was found. In some cases, the website copy was used to generate research needs.

Only documents from 2020 onwards were included, to account for the significant changes which healthcare systems have experienced as a result of COVID-19.

Documents or sections within them with a high face validity for relevant contents were read in full. Alternatively (when the table of contents and scanning the whole document did not suggest highly relevant contents, particularly when the document was not solely about healthcare, or to accommodate time constraints towards the end of the project), searches were run for *research*, R&D*, *evaluat*, gap*, knowledge, data, evidence, and method** using the "Find" function of a particular software, typically Adobe.

In the case of funding calls and funded projects – types of key stakeholder sources which differed from the rest in their structure – we reviewed website information on funding calls, lists of funded projects and, at times, funder reports.

UNDER REVIEW

*Pre-existing standalone lists of research needs*

Lists known to the team from before the start of the study were used: of the *James Lind Alliance* on *Greener Operations [11]* and of the *Royal Netherlands Academy of Arts and Sciences (KNAW) –* a *Longlist of knowledge gaps in Planetary Health [12]*. The literature searching processes for both systematic reviews and key stakeholder documents were expected to capture further standalone lists of research needs, if available.

UNDER REVIEW

#### B. SOURCES USED to generate research questions, by type

##### Systematic reviews from which research questions were derived (by chronology of data extraction):

***Targeted work on identifying knowledge gaps/ research priorities from which research questions were derived:***

11. James Lind Alliance. Greener Operations: Sustainable Peri-Operative Practice. Priority Setting Partnership Workshop Outcomes, Jun 2022. Last accessed Aug 2025.
12. Royal Netherlands Academy of Arts and Sciences (KNAW). Longlist of knowledge gaps in Planetary Health. Appendix to the report “Planetary Health. An emerging field to be developed”, 2023. Last accessed Aug 2025.

***Key stakeholder reports (13-20) and web sources (21-24) from which research questions were derived:***

13. Climate and Health Coalition. Driving Co-benefits for Climate and Health. 2022 Update: How the private sector can accelerate progress. Guidance for businesses, investors and policy makers, Nov 2022. Last accessed Aug 2025.
14. Health Care Without Harm, in collaboration with Arup. Global Road Map for Health Care Decarbonization: A navigational tool for achieving zero emissions with climate resilience and health equity. Health Care Without Harm Climate-Smart Health Care Series. Green Paper Number Two, Apr 2021. Last accessed Aug 2025.
15. NHS England. Delivering a ‘Net Zero’ National Health Service, Jul 2022.
16. NICE (National Institute for Health and Care Excellence). NICE Listens environmental sustainability project recommendations. Appendix to NICE Listens: Public dialogue on environmental sustainability. Final report, Feb 2023. https://www.nice.org.uk/what-nice-does/our-research-work/nice-listens. Last accessed Aug 2025.
17. Scottish Government, NHS Scotland. Climate Emergency & Sustainability Strategy 2022-2026, Aug 2022. Last accessed Aug 2025.

UNDER REVIEW

18. The Carbon Trust and GIG Cymru Partneriaeth Cydwasanaethau (NHS Wales Shared Services Partnership). NHS Wales Decarbonisation Strategic Delivery Plan 2021-2030, Mar 2021. Last accessed Aug 2025.
19. UK Health Alliance for Climate Change. End of Year Report 2023, Jan 2024. Last accessed Aug 2025.
20. World Health Organization. Operational framework for building climate resilient and low carbon health systems, Nov 2023. Last accessed Aug 2025.
21. Webpages of the Health for Extinction Rebellion: https://healthforxr.com/in-support-of-health-activism/. Last accessed Aug 2025.
22. Webpages of the Centre for Sustainable Healthcare, case studies: http://networks.sustainablehealthcare.org.uk/CaseStudies. Last accessed Aug 2025.
23. Webpages of NHS England. Greener NHS. System progress, case studies: https://www.england.nhs.uk/greenernhs/whats-already-happening/. Last accessed Aug 2025.
24. Webpages of the Sustainable Healthcare Coalition, on carbon calculators: https://shcoalition.org/care-pathway-carbon-calculator-2/ https://shcoalition.org/coming-soon/. Last accessed Aug 2024 (no longer live Aug 2025).

***Other stakeholders' reports from which research questions were generated:***

25. The Physiological Society and Faculty of Public Health. Red Alert: Developing a human-centred national Heat Resilience Strategy, Nov 2023. Last accessed Aug 2025.

***Funder websites reviewed in detail:***

Over 430 funding areas were screened (most on the UKRI website), the majority *not* addressing decarbonisation. Research questions were generated from 2 calls and 2 recently funded projects calls. A number of further calls were used to generate placeholder themes.

National Institute for Health and Care Research (NIHR)
Wellcome
- Climate and Health funding opportunities
- Climate and Health awarded grants

UK Research and Innovation (UKRI)
- Areas of investment and support
- Funding finder

Advanced Research + Invention Agency (ARIA)

UNDER REVIEW

Programmes

Opportunity seeds

***Background sources used to generate 'placeholder themes' and test the emerging framework*** *(organised by reason for exclusion from the pool of sources used to generate research questions):*

*Dated but highly relevant systematic reviews:*

1. Nichols A, Maynard V, Goodman B, Richardson J. Health, Climate Change and Sustainability: A Systematic Review and Thematic Analysis of the Literature. Environ Health Insights 2009;3:63-88. doi: 10.4137/ehi.s3003
2. McGain F, Naylor C. Environmental sustainability in hospitals - a systematic review and research agenda. J Health Serv Res Policy 2014;19(4):245-52. doi: 10.1177/1355819614534836

*Medical specialty-specific systematic reviews:*

*Key stakeholder publications which did not yield research questions:*

8. Aumônier S and Collins M for the Sustainable Healthcare Coalition. Healthcare, Circular Economy Principles and Sustainable Wellbeing. Undated, upload Feb 2020. Last accessed Aug 2025.
9. BMJ and UK Health Alliance on Climate Change. Net Zero Clinical Care 2023: Key Summaries Report, Jan 2024. Last accessed Aug 2025.

UNDER REVIEW

10. Care Quality Commission. “Environmental sustainability – sustainable development” element of the Single assessment framework of the CQC. Last accessed Aug 2025.
11. McGeoch, Hardie T, Coxon C and Cameron G for the Health Foundation. Net zero care: what will it take? Sep 2023. Last accessed Aug 2025.
12. Smith R, Stancliffe, R, Clark W, et al. for the UK Health Alliance on Climate Change, Centre for Sustainable Healthcare, Healthcare Without Harm Europe, Sustainable Healthcare Coalition and Health Declares Climate & Ecological Emergency. Six steps to promote recovery of the health and social care system from the covid-19 pandemic. The BMJ Opinion, 24 Sep 2020.
13. UK Health Alliance on Climate Change. Our manifesto for the UK General Election: Five priorities to sustain our health, health and care services, and environment, 2024. Last accessed Aug 2025.
14. WHO. WHO guidance for climate-resilient and environmentally sustainable health care facilities, Oct 2020. Last accessed Aug 2025.

*Key stakeholder websites which did not yield relevant documents, but elements of whose contents were used to generate placeholder themes:*

15. Greener Practice: The UK’s primary care sustainability network [website]. Last accessed Aug 2025.
16. Applied Research Collaborations (ARCs). Webpages listed under: https://www.nihr.ac.uk/about-us/what-we-do/infrastructure/applied-research-collaborations. Last accessed Aug 2025.
17. HealthTech Research Centres (HRCs). Webpages listed under: https://www.nihr.ac.uk/about-us/what-we-do/infrastructure/healthtech-research-centres. Last accessed Aug 2025.
18. Health Innovation Network [website]. Last accessed Aug 2025.

*Sources of stakeholders which were excluded from the list of “key stakeholders” once its inclusion criteria were finalised:*

19. Sawyer, M. SEE Sustainability. Climate emergency declaration: A guide for primary care, 2021. Last accessed Aug 2025.
20. SEE Sustainability. General practice non-clinical carbon calculator. Last accessed Aug 2025.

*Internal University of Cambridge sources (study was co-funded by the University of Cambridge):*

21. *University of Cambridge.* Cambridge Zero’s Climate Change Research Map. Excel spreadsheet, 2024.

UNDER REVIEW

### Supplement 2: Analysis approach – content types, transformations and principles underpinning the development of the organising framework

UNDER REVIEW

This supplement describes the process of generating research questions from the sources listed in Supplement 1. Research questions could rarely be directly extracted. Instead, they were derived through an interpretative process. Table 2a lists the types of contents used in this process. Table 2b described the types of transformations to which the source content was subjected. The section after describes the 10 principles underpinning the development and refinement of the organising framework.

**Table S2a: Types of content from which questions could be generated**

| TYPES OF CONTENTS IN SOURCE DOCUMENTS | DESCRIPTION |
|---|---|
| **Contents found across source types (systematic reviews and key stakeholder sources)** | |
| **Research gaps, needs or recommendations** | Statements which explicitly identify a research gap or need or make an explicit recommendation for a type of research to be conducted or an area of research to be prioritised.<br><br>In some cases, the sources we reviewed (systematic reviews or key stakeholder documents) pointed to a research gap or recommendation as expressed in one of the references they themselves had consulted. When there was a clear singular attribution to an external source, we reference both the source in our sample and the respective original source without, however, having accessed the latter. |
| **Research limitations** | In a systematic review, contents of this type are typically:<br>• statements from the section describing the limitations of the research, whether of the systematic review itself or of the source studies;<br>• statements about risk of bias assessments. |

| TYPES OF CONTENTS IN SOURCE DOCUMENTS | DESCRIPTION |
|---|---|
| | In policy documents, contents of this type are typically statements that describe limitations of national data. |
| **Research availability** | Statements that certain topics have been researched to a limited degree unaccompanied by an explicit research recommendation. For instance, a systematic review may state that only two studies address a particular issue yet not suggest that further research on that issue is needed.<br><br>On the one hand, the research recommendation may be considered implied, at least in a broader list of research recommendations than the one included in the review. On the other hand, an argument can be made that even if the authors identified a research gap, they made a judgement not to prioritise it and that this judgement would apply even if they could broaden their list of research recommendations. |
| **Contents found in key stakeholder sources only** | |
| **Research-policy-healthcare-frontline collaboration** | Statements which refer to an interaction between researchers/ research institutions, policy makers and practitioners, with the interaction either 'ongoing' (currently underway) or 'intended' (envisaged as future work). |
| **Recommendation for evidence-based guidance development** | Statements concerning the development of evidence-based guidance in healthcare. These have been individuated as a separate category as guidance development tends to be underpinned by an infrastructure of close links between policy, practice and academic research. |
| **"Language acts" with a non-trivial new knowledge component** | Statements in key stakeholder documents which function as verbal actions (such as a promise or a requirement) supposed to be followed by practical actions and which involve research, data, evaluation, or other form of new knowledge. The new knowledge is non-trivial and needs to be generated or shared by specialists (researchers, analysts, evaluators, domain experts), whether in thematic or methodological areas.<br><br>Examples of such language acts are directives, commitments, intentions, or acknowledgements of need. |

UNDER REVIEW

| TYPES OF CONTENTS IN SOURCE DOCUMENTS | DESCRIPTION |
|---|---|
| **Intention to fund or commission research – problem statement** | In the case of funding organisation sources, used for statements which articulate:<br>• a problem or research area which is targeted by a current funding initiative/ call for proposals. The descriptions can be quite broad but also quite specific and technical;<br>• research question(s) or research area, as per the funding proposal of the researcher or team which received the funding. Such work may be in the future or ongoing.<br><br>In the case of other key stakeholder documents, used for statements expressing an intention or recommendation to commission a study, with no specific detail of a funding call. |

UNDER REVIEW

**Table S2b: Types of transformations of source contents (used when research questions could not be directly extracted)**

| TYPE OF TRANSFORMATION | DESCRIPTION |
|---|---|
| **Decarbonisation focus attributed** | The research need was formulated in terms of healthcare 'decarbonisation' unlike the source statement which could be framed in terms of climate change, sustainability (environmental or broader) or 'being green' in the context of healthcare and/or in interaction with health outcomes.<br><br>Overall, the assumption is that the decarbonisation aspect is entailed in all other concepts, since they are broader than the concept of decarbonisation. It is nonetheless logically possible that while there is a research need at the higher level (e.g. climate change and health), it has been addressed at the lower level (decarbonisation). |
| **Boundaries of setting (re) specified** | Used when the research need was formulated about the health system (the focus of the current study) or a 'healthcare context' (underspecified), while the setting referred to in the original study was different. It could be broader than that of the health system, e.g. the health sector, or narrower, e.g. hospitals.<br><br>This transposition is largely logically justified when the boundary is narrowed down (from health sector to healthcare system) and logically problematic when the boundary is expanded (from, for instance, hospitals to health systems). In the latter case, we deemed that a question was still worth formulating, with further checks needed if the research need applies to the health system level.<br><br>Priority was given to the research topics as opposed to the settings of the studies which first proposed them. It became clear in the process of the study that most questions can be meaningfully asked for all the levels of the healthcare system. |
| **Elements individuated or re-combined** | Used when a recommendation for research includes too many elements or where related elements appear in different research recommendations. 'Elements' refers primarily to topics, but research recommendations can also specify study settings, participant types, methods, conceptual 'lenses', etc. The aim has been to keep research questions focused on one main topic, unless a judgement was made that several issues in combination make a more natural study. |
| **Claims synthesised/ level of abstraction increased** | Used when a research need has been identified in more than one source or in more than one statement within the same source. Then the phrasing and concepts from the different sources needed to be aligned and synthesised. As concepts across statements and sources are often at a different level of abstraction, such synthesis tends to mean that the resulting research need is |

UNDER REVIEW

| | |
|---|---|
| | formulated in more abstract terms than the formulation in at least one of the sources.<br><br>The more abstract concept can be from one of the statements or external to any of them (proposed by the analyst). |
| **Concept or contents articulated further** | Used when the source statement was clarified or articulated further by adding brief new contents. The goal has been to make the research question more precise, usually in cases when the source statement was not sufficiently clear. As this increases the distance from the source data, the additions are indicated in square brackets []. |
| **Research perspective added** | Used when stakeholder sources list or showcase multiple exemplars or case studies which, however, remain at the level of the particular. A research theme or question is then added proposing systematic analysis and synthesis of the case studies. |
| **Meta-science perspective added** | Used when a source statement from a key stakeholder document leads to a research question that probes something about the data collection or analysis methods referred to in the statement. (Meta-science is the practice of science using its methods upon itself, for instance to inspect its methods and improve them or to address research challenges.)<br><br>Note that this transformation is applied if the analyst made a judgement that there is a need for benchmarking against best methodological practices in academic research. If the need for scientific self-reflection is explicit in the source statement, then no such reformulation is needed. In such cases, the question is added under the *Scientific measurement and theory* category of the framework. |
| **Editing for standalone clarity** | The aim has been to formulate research questions and themes that make sense if written on a card to be discussed in stakeholder workshops or priority setting exercises. This meant that, on occasions, we needed to include contents from the broader text so that referring back to it is not needed if the questions are read without access to the sources.<br><br>Reformulating for standalone format requires primarily to:<br>• Rephrase concepts and shortcuts of expression which are only clear contextually in the source document. In some cases, this is a matter of filling back some missing words and is largely uncontroversial. In other cases, there is a need for interpretation in statement which the analyst perceives as unclear.<br>• Ensure (relative) consistency of format. Currently, the approach has been to use an explanatory phrase (representing a broad theme) for the higher- and middle-level gaps and a question format at the most specific level. In some cases, a ‘theme’ format was used at the most specific level too, as the research gap was under-specified in the source. |

UNDER REVIEW

**Main principles underpinning the development of the organising framework**

Below is a list of the ten principles applied in developing and refining the organising framework. These principles entail certain value judgements which need a broader debate. The content of the principles also needs further articulation.

1. **Variety of perspectives**

The framework recognises that there are different perspectives to the complex field it represents. It seeks to lift them into high-level categories as opposed to committing to a dominant perspective, which, in the context of decarbonisation, is typically that of sources of carbon and other greenhouse gases.

The key advantage of a multi-perspectival approach is that it enables a broad range of users to identify the perspective that best suits their organisational remit, levers of influence, priorities and mental models. Its key disadvantage is that it leads to duplication and redundancy.

2. **Some priority of the mental models of policy and decision makers**

We have given some priority to the mental models and needs of policy and decision makers, partly resulting from the orientation of the study, partly in recognition of the significant power of such stakeholders to drive change at the level of national healthcare systems.

This priority can be seen in three main choices. First, we have individuated categories for "big picture, cross-cutting issues" even if, logically, the higher-level category already captures them. For instance, there is a category of *A5.1. Transport, travel and mobility for healthcare purposes and in healthcare contexts (big picture, cross-cutting issues)* under the higher-level category of *A5. Transport, travel and mobility for healthcare purposes and in healthcare contexts.* Policy and decision makers are often interested in overviews of research, while some databases of research literature have a default setting of "automatic explosion" (automatically retrieving research on specific topics under the general topic), meaning that overview work is not easy to find if searches are run on the broad theme. Second, we created a high-level category of *Organisational levers for change*, many of which are policy-level

UNDER REVIEW

structures and mechanisms. Third, we raised into separate categories issues of trade-offs and value conflicts (see principle 7 for detail).

**3. Dynamic reformulations and re-organisation balanced against a need for standardisation and stability**

The framework was developed inductively, expanding and refining it as new research questions emerged from the source documents. We reconceptualised categories at the middle level of generality multiple times to accommodate new questions and new perspectives. We settled for the version we are presenting after we reached "light saturation" of themes: new questions continued to emerge from source documents, but they could be accommodated by existing categories. We hypothesise that the saturation we reached was sample-dependent and that broadening the inclusion criteria for document types will lead to multiple new themes, at least at the middle level of generality.

Since the field of healthcare decarbonisation is exceptionally dynamic, we argue that the framework also needs to remain dynamic and to be regularly updated to reflect new developments. However, it can serve its envisaged functions of coordinating research and action and enabling the sharing of knowledge only if stabilised in certain respects and for certain periods of time.

**4. Variety of levels of commitment to decarbonisation**

Choices about the phrasing of categories reflect an intention to remain open to different levels of endorsement of the agenda for healthcare decarbonisation, including opposition to it. On some occasions, we are explicit about the polarities of a continuum, e.g. in the phrasing of *E1.6. Legal and regulatory enablers and barriers to the decarbonisation of healthcare*. In other cases, we keep the relationship between concepts vague. For instance, the label of *D3.3. Pharmacists and the decarbonisation of healthcare* can accommodate both the action and inaction of pharmacists and various types of relationships between the two concepts and the realities behind them.

We also chose to tone down the use of strongly pro-decarbonisation language, for instance by replacing an earlier phrasing around the "*imperative* to decarbonise healthcare". Strong endorsement of the goal of decarbonisation will, undoubtedly, be dominant at the level of

UNDER REVIEW

specific sources tagged by concepts from the framework. However, we aimed to generate a framework which, at the formal level, is hospitable to all perspectives. Denial of climate change and, respectively, of the need for healthcare decarbonisation is a fundamental part of the debate which should be addressed through evidence, not conceptual barriers.

**5. User-friendliness, plain language and relative context-independence in formulations within the constraints imposed by the technical language of research**

The intention has been to avoid, as much as possible, technical language in formulating categories. For instance, we preferred a phrasing of "Healthcare decarbonisation in the social conversation" as opposed to "Healthcare decarbonisation in the social discourse".

At this stage, we have also chosen to add detail and illustrative examples in the very label of a category in order to clarify its scope as opposed to choosing briefer labels and adding scope notes. The goal has been to make a category sufficiently clear by reading its name only. For example, we have categories such as *B1. Decarbonisation of healthcare spaces (buildings, facilities, estates, sites, etc.)* and *F2.1. "Healthcare decarbonisation" – conceptualisations, boundaries with related concepts, and operationalisations* as opposed to, for instance, *Decarbonisation of healthcare spaces* and *Healthcare decarbonisation, conceptual*.

Whether the primary users of the framework would prefer detail or parsimony requires empirical testing.

Another feature of the prioritisation of user-friendliness concerned the resolution of tensions between hierarchical level in logical terms and hierarchical level in terms of visibility. When a topic received significant attention in the sources but was at a lower (less visible) hierarchical level logically, we raised it in the hierarchy to increase its visibility. This made for a worse classification system from a scientific perspective but for a more user-friendly one.

We also aimed to account for users for whom English is a second language by choosing simpler vocabulary and grammatical structures.

UNDER REVIEW

**6. Intention to align categories with relevant conceptual models**

So far, the organising framework has been developed inductively. Background knowledge of popular conceptual frameworks in healthcare (e.g. the 4 Ss of Space, Stuff, Staff and Systems) or the WHO building blocks of health systems has been used to inform some labels, but to a limited degree. We see value in incorporating, in future stages of the work, the intellectual effort accumulated in pre-existing frameworks or conceptual/ theoretical analysis.

Such frameworks may already be well standardised and embedded in classification systems used globally, e.g. the MeSH tree for types of healthcare personnel. They may be healthcare system-specific, e.g. national conceptualisations of careers in the health service, such as NHS careers. They may be associated with legal requirements, such as applying the waste hierarchy in England and Wales (DEFRA, Waste Management Plan for England, Aug 2020; p. 10). Conceptual systematic reviews, such as the Pyone et al., 2017 review of 16 frameworks of health system governance, can also enable conceptual refinements of the framework.

**7. Attention to trade-offs and value conflicts**

Complex interactions and trade-offs have been lifted to higher levels. This reflects a hypothesis that barriers to action on decarbonisation are less a matter of disagreeing about its importance than of complex, at times tragic, dilemmas of using healthcare resources and managing short-term demand vs. long-term needs.

**8. Attempt at balancing the formal, transferable and the thematic, domain-specific**

An attempt has been made to abstract formal features of approaches as opposed to prioritise their domain-belonging, even though domain-centred categories are also systematically represented. For instance, a research question around using traditional medicines as a guide in drug discovery led to the category of *Combining innovation with a 'back to basics' approach*, as opposed to only generating a drug discovery-related category.

**9. Use of the formal to pave the way to synthesis as opposed to reinforcing silos**

An attempt has been made to enable a comparative perspective and/or synthesis where contrasts and conflict seem to dominate. The example above of *Combining innovation with a*

UNDER REVIEW

*‘back to basics’ approach* is relevant here too, or the bringing together of *Top-down and/or bottom-up approaches to driving healthcare decarbonisation.*

**10. Recognition of grey zones between healthcare decarbonisation and broader climate and health issues**

Case-by-case decisions were made about remaining in decarbonisation boundaries or expanding towards climate change or sustainability. The scope is broadened when a decarbonisation focus was perceived as artificial relative to the (likely) nature of the practical work and dominant debate. For instance, some of the subthemes under *A3. Waste, reuse, recycling and circularity in healthcare* concern primarily pollution as opposed to decarbonisation. Such boundary or even out-of-scope topics can offer important opportunities to bridge debates which are distinct but also closely inter-related.

UNDER REVIEW

### Supplement 3: Full NHS-SOS framework

| ***A. NATURAL RESOURCE USE AND SOURCES OF CARBON*** |
|---|
| **A1. Energy sources and water use in healthcare** |
| A1.1. Energy sources and water use in healthcare (big picture, cross-cutting issues) |
| A1.2. Non-renewable energy sources [PLACEHOLDER] |
| A1.3. Renewable energy sources |
| A1.4. Water use |
| A1.5. Heating and cooling |
| A1.6. Lighting |
| A1.7. Energy use for new digital demands |
| **A2. Medicines, equipment and consumables as decarbonisation targets; associated production, procurement and supply chains** |
| A2.1. Medicines, equipment and consumables as decarbonisation targets; associated production, procurement and supply chains (big picture, cross-cutting issues) |
| A2.2. Medicines as decarbonisation targets |
| A2.3. Disinfecting and protective consumables and equipment as decarbonisation targets |
| A2.4. Medical gases as decarbonisation targets |
| **A3. Waste, reuse, recycling and circularity in healthcare** |
| A3.1. Management of waste and waste reduction (big picture, cross-cutting issues) |
| A3.2. Innovations in waste management |
| A3.3. Reducing waste |
| A3.4. Reuse |
| A3.5. Recycling |
| A3.6. Waste from healthcare – by type |
| A3.6.1. Medical and hazardous healthcare waste |
| A3.6.2. Wastewater from healthcare facilities |
| A3.6.3. Plastic waste, clinical [PLACEHOLDER] |
| **A4. Food and nutrition in healthcare contexts and for health-related outcomes** |
| A4.1. Stakeholders' knowledge, perceptions and attitudes to sustainable nutrition |
| A4.2. Polices, guidelines, strategies and plans on sustainable nutrition |
| A4.3. Networks and communities of practice for sustainable nutrition |
| A4.4. Complex interventions on sustainable nutrition [PLACEHOLDER] |
| A4.5. Low-carbon nutrition in interaction with other principles of sustainable nutrition (e.g. health benefits, equality, affordability) [PLACEHOLDER] |
| A4.6. Shaping sustainable eating behaviours of patients and staff |
| **A5. Transport, travel and mobility for healthcare purposes and in healthcare contexts** |

UNDER REVIEW

| |
|---|
| A5.1. Transport, travel and mobility for healthcare purposes and in healthcare contexts (big picture, cross-cutting issues) |
| A5.2. The healthcare fleet |
| A5.3. Staff travel, transport and mobility |
| A5.4. Patient travel, transport and mobility for the purpose of accessing healthcare [PLACEHOLDER] |
| ***B. HEALTHCARE CONTEXTS*** |
| **B1. Decarbonisation of healthcare spaces (buildings, facilities, estates, sites, etc.)** |
| B1.1. Decarbonisation of healthcare spaces (big picture, cross-cutting issues) |
| B1.2. Decarbonisation of hospitals |
| B1.3. Decarbonisation of operating theatres |
| B1.4. Decarbonisation of sterilisation units |
| B1.5. Decarbonisation of chemotherapy units |
| **B2. Decarbonisation of healthcare settings (primary care, secondary care, community care, etc.) [PLACEHOLDER]** |
| **B3. Decarbonisation of clinical specialities and/or health conditions** |
| B3.1. Surgery as a decarbonisation target |
| B3.2. Asthma care as a decarbonisation target [PLACEHOLDER] |
| B3.3. Radiology and radiotherapy as decarbonisaton targets |
| B3.4. Obstetrics and gynaecology as decarbonisation targets |
| B3.5. Otorhinolaryngology as a decarboisation target |
| B3.6. Dentistry as a decarbonisation target |
| **B4. Decarbonisation of healthcare supply chains** |
| **B5. Sustainable reorganisation of healthcare systems in contexts of emergencies, disasters and other stresses and shocks** |
| ***C. SOLUTIONS*** |
| **C1. Solutions for decarbonising healthcare (big picture, cross-cutting issues)** |
| **C2. Digital solutions** |
| **C3. Innovative materials** |
| **C4. Innovative designs** |
| **C5. Capture and removal of greenhouse gases [PLACEHOLDER]** |
| **C6. New models of care and decarbonisation** |
| **C7. Combining innovation and 'back to basics' approaches** |
| **C8. Improved healthcare efficiency as a route to decarbonisation** |

UNDER REVIEW

| |
|---|
| C8.1. Improved prevention |
| C8.2. Improved diagnosis |
| C8.3. Minimising the provision of care of limited or no benefit |
| C8.4. Reducing overprescribing and overuse of medicines |
| C8.5. Improved chronic disease management |
| C8.6. Improved healthcare efficiency – by clinical specialty or health condition |
| **C9. Education, training and information provision** |
| **C10. Behaviour change-focused solutions for decarbonising healthcare** |
| **C11. Complex interventions for decarbonising healthcare** |
| **C12. The carbon footprint of adaptations to climate change and decarbonisation solutions themselves** |
| C12.1. The carbon footprint of digital health |
| C12.2. The carbon footprint of increasing the climate resilience of healthcare |
| C12.3. The carbon footpring of cooling technologies in healthcare |
| ***D. STAKEHOLDERS*** |
| **D1. Stakeholders in healthcare and/or decarbonisation (big picture, cross-cutting issues)** |
| **D2. The healthcare workforce and the decarbonisation of healthcare** |
| D2.1. Healthcare workforce planning and the decarbonisation of healthcare |
| D2.2. Training, development and capacity building for the decarbonisation of healthcare |
| D2.3. Healthcare workers' behaviour change for the decarbonisation of healthcare |
| D2.4. Formal responsibilities and accountability of healthcare workers concerning the decarbonisation of healthcare |
| D2.5. Healthcare workers' activism on climate change |
| D2.6. Healthcare workers' needs for psychological support related to climate change |
| D2.7. Impact on healthcare workers of work environments responding (or not) to climate change |
| **D3. Types of healthcare workers and the decarbonisation of healthcare** |
| D3.1. Doctors and the decarbonisation of healthcare [PLACEHOLDER] |
| D3.2. Nurses and the decarbonisation of healthcare [PLACEHOLDER] |
| D3.3. Pharmacists and the decarbonisation of healthcare |
| D3.4. Allied health profesionals and the decarbonisation of health care [PLACEHOLDER] |
| **D4. Patient populations and groups and the decarbonisation of healthcare [PLACEHOLDER]** |
| ***E. ORGANISATIONAL LEVERS FOR CHANGE*** |
| **E1. Policy and governance for the decarbonisation of healthcare** |

UNDER REVIEW

| |
|---|
| E1.1. Policy and governance (big picture, cross-cutting issues) |
| E1.2. Leadership |
| E1.3. Stakeholders [See Section D] |
| E1.4. Embedding principles of healthcare decarbonisation in organisational structures and processes |
| E1.5. Top-down and/or bottom-up approaches to driving the decarbonisation of healthcare |
| E1.6. Legal and regulatory enablers and barriers to the decarbonisation of healthcare |
| E1.7. Targets, pledges and commitments to enable the decarbonisation of healthcare |
| **E2. Funding and financial mechanisms for the decarbonisation of healthcare** |
| **E3. Practical guidance for the decarbonisation of healthcare** |
| F3.1. Case studies of decarbonisation initiatives |
| F3.2. Toolkits and other ‘how to’ guides for designing and/or implementing decarbonisation initiatives |
| **E4. Certification and accreditation schemes for healthcare decarbonisation [PLACEHOLDER]** |
| **E5. Visions, models and frameworks of healthcare provision which incorporate decarbonisation principles** |
| E5.1. “Realistic medicine” (Scotland) |
| **E6. Networks, communities of practice and platforms for collaborative work on healthcare decarbonisation** |
| E6.1. Co-production and co-implementation of decarbonisation research by research, healthcare, business and/or policy actors [PLACEHOLDER] |
| E6.2. Networks within healthcare working towards decarbonisation |
| E6.3. Intersectoral collaborations for decarbonisation, with participation from the health sector [PLACEHOLDER] |
| **E7. Managing (perceived) trade-offs and conflicts of values in decarbonising healthcare** |
| E7.1. Balancing clinical effectiveness, patient preferences, financial and environmental costs |
| E7.2. Addressing the challenge of upfront investment now to achieve decarbonisation benefits later [PLACEHOLDER] |
| E7.3. Values, ethics and human rights in healthcare decarbonisation |
| E7.4. Decision making and risk management frameworks |
| ***F. SCIENTFIC MEASUREMENT AND THEORY*** |
| **F1. Measuring the carbon footprint and the decarbonisation of healthcare** |
| F1.1. Comprehensive methods and tools for calculating carbon footprint |
| F1.2. Source-of-carbon-specific methods and tools for calculating carbon footprint |

UNDER REVIEW

| |
|---|
| F1.3. Methods and tools for calculating carbon footprint as per their overall methodological-theoretical orientation |
| F1.4. Sensors and sensing systems for monitoring greenhouse gas emissions in healthcare contexts [PLACEHOLDER] |
| F1.5. Impact measures in healthcare decarbonisation |
| F1.6. Comparative measures in healthcare decarbonisation |
| F1.7. Quality standards for data, evidence and metrics on healthcare decarbonisation |
| F1.8. Integrated monitoring, reporting and evaluation |
| F1.9. Decarbonisation targets [PLACEHOLDER] |
| F1.10. Domain-specific issues in decarbonisation measurement |
| **F2. Concepts, frameworks and theories concerning the decarbonisation of healthcare** |
| F2.1. "Healthcare decarbonisation" – conceptualisations, boundaries with related concepts, and operationalisations |
| F2.2. Applied interdisciplinary fields of inquiry with applications in healthcare decarbonisation |
| F2.2.1. Decision science for healthcare decarbonisation |
| F2.2.2. Stakeholder theory for healthcare decarbonisation |
| F2.2.3. Quality improvement frameworks for healthcare decarbonisation |
| **F3. Healthcare decarbonisation in the social conversation** |
| **F4. Needs for quality improvement in research on healthcare decarbonisation** |
| **F5. Systematic approaches to identifying research and gaps in research on healthcare decarbonisation** |
| **F6. Turning knowledge into action on healthcare decarbonisation** |

UNDER REVIEW
